# Assessing the role of polygenic background on the penetrance of monogenic forms in Parkinson’s disease

**DOI:** 10.1101/2021.06.06.21253270

**Authors:** Emadeldin Hassanin, Patrick May, Rana Aldisi, Peter Krawitz, Carlo Maj, Dheeraj Reddy Bobbili

## Abstract

**Background:** Several rare and common variants are associated with Parkinson’s disease. However, there is still an incomplete penetrance in the carriers of rare variants associated with Parkinson’s disease. To address this issue, we investigated whether a PRS calculated from significant GWAS SNPs affects the penetrance of Parkinson’s disease among carriers of rare monogenic variants in known Parkinson’s disease genes and those with a family history.

**Methods:** We calculated the PRS based on common variants and selected the carriers of rare monogenic variants by using the exome data from UK Biobank. Individuals were divided into three risk categories based on PRS: low (<10%), intermediate (10%-90%), and high (>90%) risk groups. We then compared how PRS affects Parkinson’s disease risk among carriers of rare monogenic variants and those with family-history.

**Results:** We observed a two-fold higher odds ratio for a carrier of a monogenic variant that had a high PRS (OR 4.07,95% CI, 1.72-8.08) compared to carriers with a low PRS (OR 1.91, 95% CI, 0.31-6.05). In the same line, carriers with a first-degree family history and with >90% PRS have even a higher risk of developing PD (OR 23.53, 95%CI 5.39-71.54) compared to those with <90% PRS (OR 9.54, 95% CI 3.32-21.65).

**Conclusions:** Our results show that PRS, carrier status, and family history contribute independently and additively to the Parkinson’s disease risk.

## Introduction

Parkinson’s disease (PD) is a complex disorder where genetics, including rare and common variants, plays a crucial role in the disease etiology^1^. During the past years, several rare and highly damaging variants were associated with PD via familial and case-control studies^2–5^. In addition to the rare risk variants, common variants also contribute to PD development^6^. Genome-wide association studies (GWAS) have identified an increasing number of PD risk loci^7,8^. Alone, each of these common variants has relatively modest effect compared to rare risk variants on the overall disease risk for PD. However, when combined in the form of polygenic risk scores (PRS), common risk factors confer significantly high disease risk^9^. PRS are generated per individual, and notably, when comparing those individuals within the highest percentile with the rest of the population, the effect is almost at the same level as those with monogenic variants^10^. PRS have recently gained traction as one of the critical risk stratification tools in PD and other complex diseases^11,12^. In PD, a considerable amount of work has been done on PRSs and their effect on the disease status^13^, age of onset^14^, and carrier status^15^. In addition to being a risk factor by itself, in the previous studies, the effect of PRS on the penetrance of damaging *GBA*^16^, and *LRRK2* p-G2019S (rs34637584) variants has been studied^15^. A recent GWAS has identified a significant locus in the *CROC1* gene associated with the penetrance of *LRRK2* variants^17^. However, previous studies have focused only on particular genes and variants, and a study to measure the effect of PRS on the other genes and variants is still missing. Several studies have been performed in the same lines to study the impact of rare damaging heterozygous variants in the canonical PD genes^18^. Despite this, a clear association has not been established between rare heterozygous variants and PD. Hence, it is highly crucial to study the effect of PRS on the penetrance of rare heterozygous variants in all the monogenic forms of PD, which is mainly hampered by the lack of large enough cohorts.

One factor that can explain the incomplete penetrance in the carriers of rare heterozygous variants is the polygenic background, as shown previously for *LRRK2* G2019S carriers^15^. As a continuation to the earlier studies, for the first time in PD, we studied the effect of PRS on the carriers of rare PD risk variants in those genes that were previously identified as associated with PD. To do this, we used the whole exome data (WES) from ∼200K individuals from UK Biobank (UKB) data. Further, we examined the combined impact of rare damaging heterozygous PD risk variants, PRS, and family history on PD development. According to our knowledge, none of the previous studies have performed such an integrated analysis in the known PD genes.

## Material and methods

### Data Source

This study was performed using genotypic and phenotypic data from the UKB application number 43140 and Parkinson’s progression markers initiative (PPMI) data^19^. UKB is a long-term prospective study, and volunteers are being recruited mainly from England, Scotland, and Wales, and involves more than 500,000 participants aged between 40-69 years, with 200,643 exomes sequenced. The dataset is available for research purposes, and all participants provided documented consent^20^. PPMI is an ongoing multicentre prospective cohort study with a longitudinal follow-up. PPMI aims to identify blood-based, genetic, spinal fluid, and imaging biomarkers of Parkinson’s disease progression^21^. We used the genotype and WES data from the PPMI.

### Study cohort

Diagnosis of PD was based on self-reported illness codes, the International Classification of Diseases (ICD-10) diagnosis codes. This included individuals with self-reported code 1262, or ICD-10 code of G20 in hospitalization records. The quality control for both genotype and WES has been performed by the UKbiobank and we used the processed files in the downstream analysis. We included individuals of all ancestries. We excluded outliers with putative sex chromosome aneuploidy, high heterozygosity or missing genotype rates, and with discordant reported versus genotypic sex. The outliers are defined as those coded as “YES” in the fields 22019, 22027, and 22001 respectively. Only individuals with both genotyping and WES data were included. Analysis was restricted to unrelated individuals up to second-degree. We excluded one from each pair of related individuals if the genetic relationship was closer than the second degree, defined as kinship coefficient > 0.0884 as calculated by the UK Biobank. The PPMI data were filtered according to the procedures followed in the previous study^9^.

### Exome data filtering

#### Annotation

Annotation for the WES data was performed using ANNOVAR^22^ against the hg38 human reference genome version with default parameters. The Genome Aggregation Database (gnomAD)^23^, was used to retrieve variant frequencies, and ClinVar^24^ annotations were used to interpret the pathogenicity of germline variants.

#### Variant selection

The following criteria were employed to select the heterozygous, rare damaging variants (VARrd) from UK biobank data. 1) Minor allele frequency < 0.005 based on gnomAD, 2) not annotated as “synonymous,” “non-frame shift deletion,” and “non-frameshift insertion,” 3) annotated as “pathogenic” or “likely pathogenic” based on ClinVar and 4) annotated as associated to “Parkinson’s disease” based on ClinVar. From the ClinVar dataset we excluded TNK2 gene as it is not commonly considered as PD gene and the ClinVar annotation for it was based on previous whole-exome studies^25^ with no further experimental evidence that confirmed the pathogenic role of this gene in PD. In the next step, we extracted the VARrd from the WES vcf file to identify the carriers and non-carriers using PLINK2^26^.

### Polygenic risk score analysis

To generate the PRS, we used the summary statistics from a list of 90 SNPs discovered from the most extensive available meta-analysis of PD to date^8^. To ensure that we are not counting the variants twice in both carrier status and PRS calculation, we excluded the VARrd present in the 90 GWAS SNPs. Only LRRK2 G2019S was common between the two sets. Hence, we proceeded with the remaining 89 SNPs for PRS calculation using PRsice2^27^ (Supplementary Figure S1), while accounting for allele-flipping and the removal of ambiguous SNPs automatically. We used the ‘–no-regress’ and ‘–no-clumping’ options along with default parameters. One of our goals in this study is to include as many ancestries as possible. To do that, an adjusted PRS was generated using a previously defined approach to reduce variation in PRS distribution through genetic ancestry^28^. As a first step, we used the first four principal components of ancestry, provided by UKB^29^, to fit a linear regression model. Using that model (PRS∼PC1 + PC2 + PC3 + PC4), we predicted the ancestry PRS in the entire dataset. We then subtract this predicted ancestry PRS from the raw PRS to calculate a residualized score. A residualized score (ancestry adjusted PRS) was used to categorize individuals (Supplementary Figure S2). With this approach, we aimed to address the transferability of PRS in other ancestries similar to a previous study^30^.

### Statistical analysis

The next step after generating the PRS was to stratify the individuals belonging to different PRS percentiles. Here, we used the *ntile* function in the R package “dplyr” to divide the individuals into percentile groups. We assigned the low, Intermediate, and high labels based on the percentiles the individuals belong to 1-10% (low), 10-90% (intermediate), and 90-100% (high). We chose the 90-100% percentile as the “high” category as it showed the highest effect size when we compared the different percentiles as shown in Supplementary Figure S3.

To compare the effect of PRS on carriers or non-carriers of VARrd, we used the non-carriers labeled as “intermediate” as the reference group. We compared the carrier and non-carriers in the high-PRS group against the low-PRS group using a logistic regression model while adjusting for age at study, sex, and the first four principal components. We then measured the impact of PRS on carriers in each gene, and individuals with variants in more than one gene were excluded.

To quantify the combined effect between VARrd, family history of PD, and development of PD, we calculated the odds ratios of PD occurrence in those with a family history of PD and those without for the different groups based on PRS percentiles. Here, we only used two PRS categories non-high (<90%) and high (90%). Such classification has been performed because there are very few individuals with family history in the low category leading to biased estimates. For all statistical analyses, we used R 3.6.3.

## Results

### UK biobank dataset description

Since we were using both common and rare exonic variants in our analysis, we restricted our analysis to the UKBB 193,330 individuals for which WES and genotype data were available. Descriptive statistics of the study population after the filtering steps are shown in Table 1 and Supplementary Table S1. In total, we identified 806 cases with PD with the self-reported mean age at PD onset of 64.58 years. 59.43% of PD cases were male. We considered 192,524 participants as controls with a mean age at recruitment of 56.44 years. We identified 2,980 carriers of 43 pathogenic or likely pathogenic variants related to PD in 17 genes associated with PD (Supplementary file 1). In PPMI, we identified 15 carriers with VARrd in three genes (*LRRK2, GBA, PRKN*). We provided the list of all the VARrd along with their annotation in Supplementary file 1. The distribution of age of onset in the carriers of VARrd is shown in Supplementary Figure S4.

**Table 1:** Characteristics of the 193,330 UK Biobank participants by Parkinson’s disease (PD) status.

|  | **Cases** | **Controls** |
| --- | --- | --- |
| **Participants, N** | 806 | 192524 |
| **Male, N (%)** | 479 (59.43) | 86556 (44.96) |
| **Female, N (%)** | 327 (40.57) | 105968 (55.04) |
| **Age\*, mean (SD)** | 64.58 (8.59) | 56.44 (8.08) |
| **Family history of parkinsons's disease, n (%)** | 93 (11.54) | 8011 (4.16) |

\*Age: age at onset for cases, and age at last visit for controls.

**Table 2:** Characteristics of the 486 PPMI participants.

|  | **Cases** | **Controls** |
| --- | --- | --- |
| **Participants, N** | 341 | 145 |
| **Male, N (%)** | 223 (65.4) | 94 (64.83) |
| **Female, N (%)** | 114 (33.43) | 50 (34.48) |
| **Age, mean (SD)** | 61.8 (9.55) | 60.94 (10.46) |
| **Family history of parkinsons's disease, n (%)** | 87 (25.51) | 7 (4.83) |

### Polygenic background modifies the risk in monogenic variant carriers

We investigated how polygenic risk alters the penetrance of the variant carriers in PD. We used PRS to categorize individuals into low (1-10%), intermediate (10-90%), and high risk (90-100%) groups based on PRS percentiles. We then compared how the PRS among carriers influences the PD risk. Compared to non-carriers with low PRS, PD risk among carriers with low PRS was (OR=1.91, 95% CI, 0.31-6.05), which increased to (OR=4.07, 95% CI, 1.72-8.08, p=3e−04) among carriers with high PRS (Figure 1).

**Figure 1:**
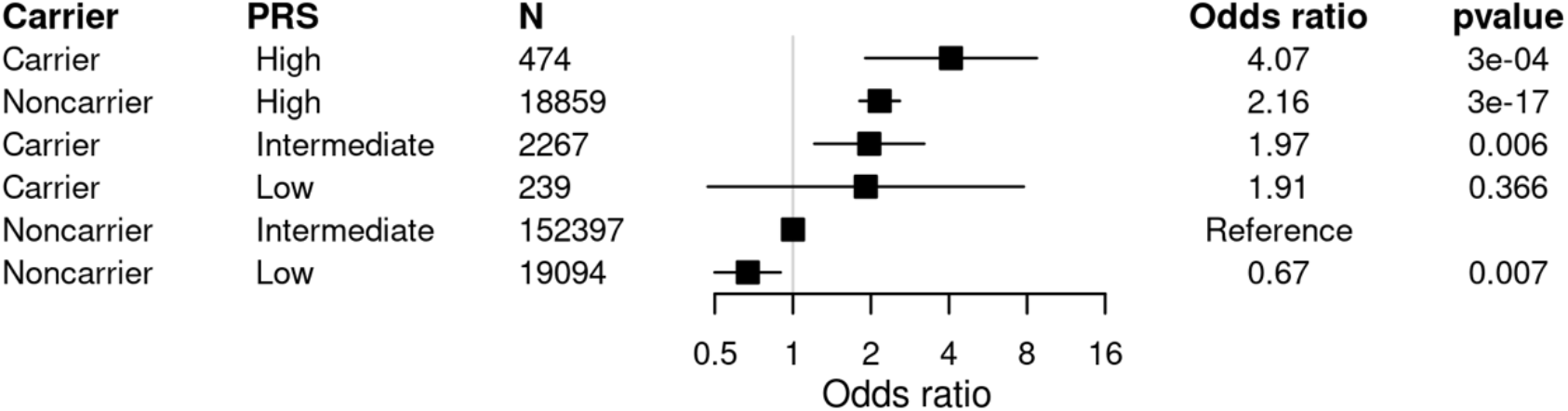
Impact of polygenic risk score (PRS) across carrier and non-carrier of monogenic variants in the UK Biobank cohort. Carriers and non-carriers were categorised into three groups based on their PRS percentiles: low (<10%), Intermediate (10-90%) or high (>90%). The odds ratio (OR) was calculated from a logistic regression model with age, sex, and the first four principal components of ancestry as covariates. The reference group was individuals with no qualifying variants (NonCarriers) with intermediate PRS. The adjusted ORs are indicated by the black boxes. The 95% confidence intervals are indicated by the horizontal lines around the black boxes.

### The impact of polygenic risk in *LRRK2, GBA*, and *PRKN* variant carriers

Only three genes, *LRRK2, GBA*, and *PRKN*, out of 17 PD-associated genes had qualifying variants in cases. There were no VARrd in cases in the remaining 14 genes. The number of carriers for each variant and gene is given in supplementary file 1. Therefore, we proceeded with those three genes and estimated how the PRS affects PD risk in *LRRK2, GBA*, and *PRKN* VARrd carriers. The distribution of PRS in *LRRK2, GBA*, and *PRKN* VARrd can be seen in Supplementary Figure S5. Since there were no cases with *LRRK2* and *GBA* variants and only two cases with *PRKN* variants in the low PRS group, we stratified samples into only two groups according to PRS percentiles: non-high (<90%) and high (>90 %). We compared high PRS and non-high PRS groups for variant carriers versus non-carriers for each gene independently. For carriers with variants in *LRRK2* and *GBA*, a high PRS further increased the PD risk. Compared to cases with no *LRRK2* variants, *LRRK2* variant carriers had an OR of 55.66 (95% CI 11.94–194.12) for the high PRS group and an OR of 8.10 (95% CI 2.82–18.33) for the non-high PRS group. *GBA* variant carriers in the low PRS groups had an OR of 1.77 (95% CI 0.44–4.68), which increased to 3.58 (95% CI 0.88–9.62) in those with non-high PRS. *PRKN* carriers in the non-high PRS group had an OR of 1.84 (95% CI 0.91–3.27), which slightly decreased to 1.58 (95% CI 0.09–7.16) in *PRKN* carriers with high PRS (Supplementary Figure S6).

### The impact of polygenic risk in individuals with first-degree family history

We first estimated the effect of first-degree family history in developing PD by comparing carriers and non-carriers of VARrd with and without family history. It can be clearly seen that family history itself has an independent effect on developing PD as seen in Supplementary Figure S7. Furthermore, we then compared how PRS stratification affects the PD risk among individuals with first-degree family history. The OR for PD in individuals with first-degree family history - as compared to non-familial cases in the intermediate PRS group - was 2.21 (CI% 0.87-4.55) in the low PRS group and 5.30 (CI% 3.33-8.01) in the high PRS group (Figure 2).

**Figure 2:**
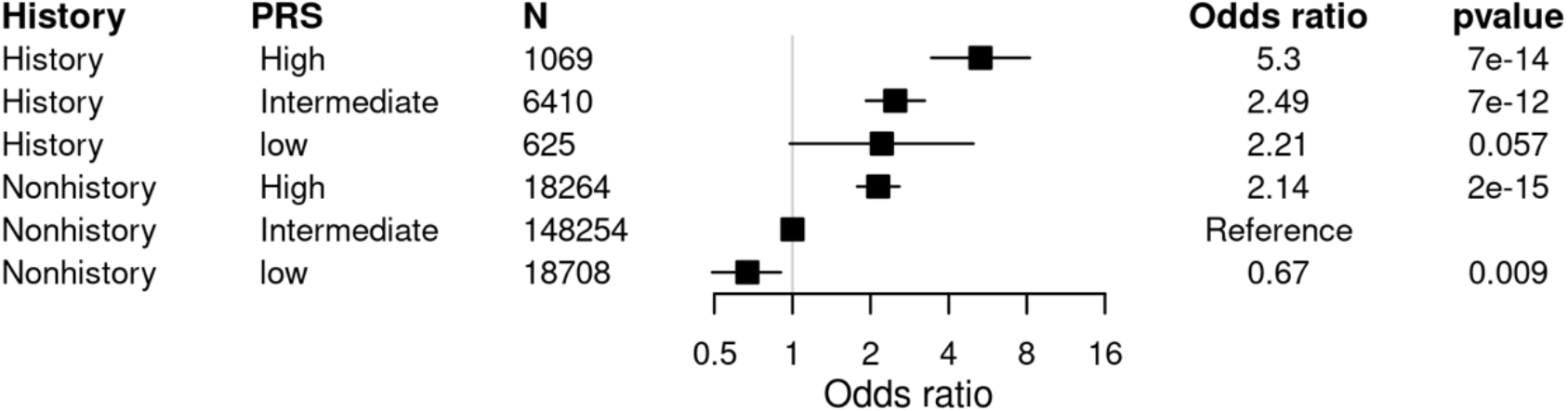
Impact of the polygenic risk score (PRS) across individuals with a first-degree family history diagnosed with Parkinson’s disease (PD) in the UK Biobank cohort. PRS was categorised into three groups based on the percentiles: low (<10%), Intermediate (10-90%) or high (>90%). The odds ratio (OR) was calculated from a logistic regression model with age, sex, and the first four principal components of ancestry as covariates. The reference group was individuals with no family history (Nonhistory) with intermediate PRS. The adjusted ORs are indicated by the black boxes. The 95% confidence intervals are indicated by the horizontal lines around the black boxes.

We next examined whether PRS stratification affects the PD risk among variant carriers with a first-degree family history. Again, we used PRS to categorize individuals into two groups: non-high (<90%) and high risk (>90) groups. The OR for PD in carriers with first-degree family history - as compared to non-carrier with no PD history - ranged from 9.54 (CI% 3.32-21.65) for individuals in the non-high PRS group to 23.53 (CI% 5.39-71.54) in the high PRS group (Figure 3).

**Figure 3:**
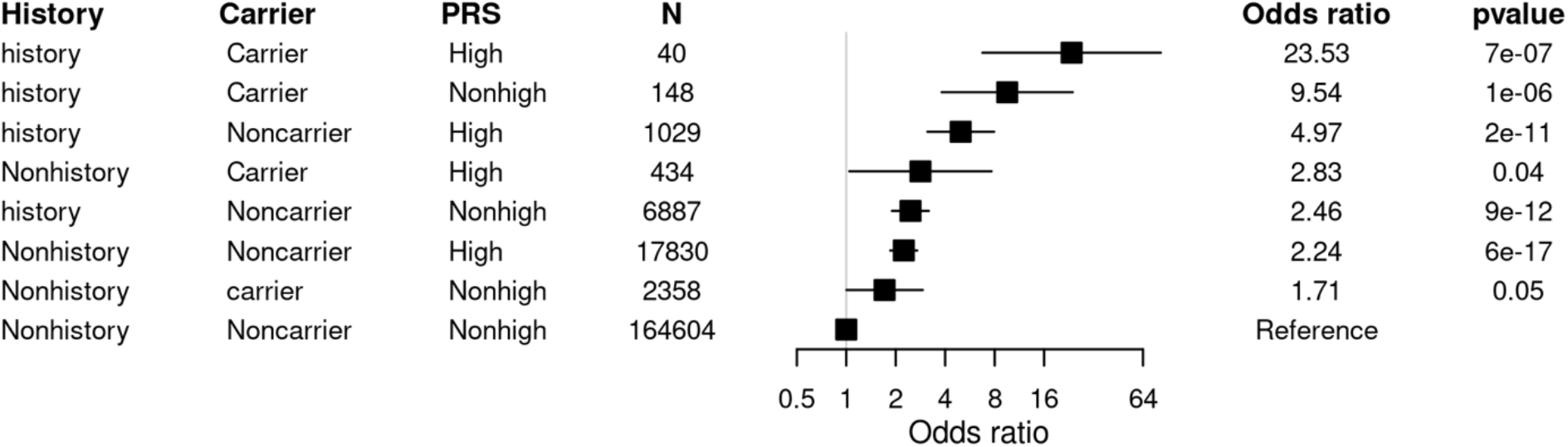
Impact of the polygenic risk score (PRS) across individuals with both monogenic variants and a first-degree family history diagnosed with Parkinson’s disease in the UK Biobank cohort. Carriers and non-carriers with and without having a first-degree family history were categorised into two strata based on their PRS percentile: Non-high (<90%), or high (>90%). The reference group was individuals with no qualifying variants (NonCarriers) and no family-history and non-high PRS. The adjusted odds ratios (ORs) are indicated by the black boxes. The 95% confidence intervals are indicated by the horizontal lines around the black boxes.

### Validation in the PPMI cohort

We explored the WES and genotyping dataset from the Parkinson’s progression markers initiative (PPMI) for validation. We could not use the PPMI cohort for risk stratification because it contained only 15 PD risk variant carriers. However, in the PPMI dataset, we could explore the number of cases in each PRS strata. There was one case among the variant carriers in the low PRS and six in the high PRS group, respectively (Supplementary Table S2). We then explored whether the polygenic background modifies the PD risk conferred by positive first-degree family history in the PPMI cohort. We then investigated whether PRS affects the PD risk conferred by positive first-degree family history in the PPMI cohort. All individuals with a family history of PD and in the top PRS decile were cases (Supplementary Table S3).

## Discussion

The results presented in this paper show that the PRS is significantly associated with the penetrance of rare damaging variants in PD. Here, we demonstrated that risk conferred by monogenic risk variants could be affected considerably by PRS, which is assumed to act by affecting multiple pathways related to PD^31^. Our study further supports the findings from another study conducted on *LRRK2* p.G2019S^15^. These results are broadly consistent with a liability threshold model for PRS. The probability that any given pathogenic variant carrier crosses the threshold to develop a disease is influenced by the underlying liability conferred by the polygenic background^32^. Few pharmaceutical companies are conducting clinical trials by recruiting only PD cases with monogenic variants in *LRRK2* or *GBA* into account^33^. Hence, it is imperative to understand the combined effect of PRS and those monogenic risk variants for choosing the trial participants and designing personalized therapeutic strategies. Family history is known to be a significant risk factor in PD. However, we show for the first time that there is almost a two-fold increase in PD risk in those individuals that have both a family history of PD and high PRS (P-value=7e−14, OR=5.30, CI=3.33-8.01) compared to those with family history and low PRS (P-value=0.057, OR=2.21, CI=0.87-4.55) (Figure 2) and this effect is even more pronounced when taking carrier status into account (P-value=7e−07, OR=23.53, CI=5.39-71.54) (Figure 3). These findings highlight that it might be worth considering PRS while making clinical decisions in both carriers and non-carriers of VARrd. The current standard practice to assess rare monogenic risk variants in a clinical setting is to perform gene panel sequencing, clinical-grade WES, or whole-genome sequencing (WGS). However, only WGS can take both monogenic and common variants into account of the technologies mentioned above. Nevertheless, comparatively, it is at least 2-3 times more expensive than WES. Hence, it might be worth routinely generating genotype data in addition to WES or targeted gene sequencing to capture the common variants additionally to be able to calculate the PRS. This will significantly improve the ongoing efforts for saving health care costs and will increase the accessibility for larger cohorts for which both rare variant and PRS will be available. It also enhances the feasibility of incorporating PRS information into standard clinical practice. Further, we need to develop multifactorial models integrating genetic and non-genetic risk factors similar to those that are routinely being used such as BOADICEA model in the breast cancer^12^.

This study of analyzing PD prevalence in the UKBB cohort with exome and genotyping data shows the independent role of each polygenic risk, family history, and high-impact genetic variants on the PD risk. The polygenic risk strongly alters the incidence of PD not only in sporadic cases but also in monogenic forms. Our analysis also suggests that family history for PD represents an additional risk factor to the genetic data and contributes independently to the disease risk. Thus, PRS, rare high-impact variants, and family history act independently and additively on the PD risk. These findings highlight the potential of considering PRS in the clinical risk stratification of PD.

There are several limitations to this study. The major one being the lack of a large enough replication dataset. Currently, not many datasets with common and rare variant data are available. We explored the WES and genotyping dataset from PPMI. However, there were only 15 carriers of monogenic variants, and hence we could not use that dataset for full replication. Although we replicated a few findings from previous studies for either common variants or single genes and showed that our results align with the field’s current knowledge, it would be essential to replicate our research findings independently.

Another limitation is that the UKbiobank is not aimed only towards PD alone. Hence, the age and sex distribution are not uniform in the current study. We addressed this problem by using the age and gender as covariates. However, future studies should address this issue, and these findings should be replicated in well-balanced PD cohorts^34^. The sample size is another limitation of our study. Even though it is one of the most extensive WES studies in PD so far, while categorizing the individuals as carriers and non-carriers, some of the categories have very few individuals leading to large confidence intervals. Hence, some of the effect sizes may be overestimated in those categories. It would be necessary for the future to include more and better phenotyped individuals in the analysis to perform similar studies.

Our study aimed to be inclusive and had all the study individuals in the investigations identical to a previous study^28^. One drawback of this approach is that there could be population stratification due to various ethnicities, and the current dataset is dominated by “White British” individuals. Although we tried to provide a possible solution to this problem by exploring the possibility of using ancestry-adjusted PRS in all ethnicities^29^, the current findings need to be validated in specific ethnicity-controlled cohorts. The population bias is a significant limitation in our research and across most genetic studies. Hence, it is crucial to include multiple ethnicities in future studies to understand and transfer research findings. In our study, we classified variants based on ClinVar annotation alone. In clinical practice, geneticists follow much stricter guidelines such as the ACMG classification^35^ in prioritizing the variants. We aimed to perform an unsupervised approach, and hence we restricted our analysis to only using ClinVar as the source for pathogenic PD variants. Overall, our study highlights the contribution of PRS in developing PD and provides a comprehensive evidence that PRS can be an effective tool in the disease risk stratification. Especially, in those individuals that are carriers of rare variants. In combination with the other risk factors, it can be a cost-effective addition in identifying individuals with high and low risk of PD and aid clinicians in taking necessary actions.

## Data Availability

Genome-wide genotyping data, exome-sequencing data, and phenotypic data from the UK Biobank and Parkinson's Progression Markers Initiative (PPMI) are available upon successful project application.

https://www.ukbiobank.ac.uk/

https://www.ppmi-info.org/

## Acknowledgments

UK Biobank analyses were conducted via application 43140 using a protocol approved by the Partners HealthCare Institutional Review Board. Data used in the preparation of this article were obtained from the Parkinson’s Progression Markers Initiative (PPMI) database (www.ppmiinfo.org/data). For up-to-date information on the study, visit www.ppmiinfo.org.

## Author Roles

(1) Research Project: A. Conception, B. Organization, C. Execution; (2) Statistical Analysis: A. Design, B. Execution, C. Review and Critique; (3) Manuscript Preparation: A. Writing of the First Draft, B. Review and Critique.

E.H.: 1A, 1B, 1C, 2A, 2B, 3A

P.M.: 1A, 1B, 1C, 2C, 3B

R.A.: 2C, 3B

P.K.: 2C, 3B

C.M.: 1A, 1B,1C, 2A, 2C, 3B

D.R.B.: 1A, 1B, 1C, 2A, 2C, 3A, 3B

