## Supplementary_material for "Assessing the role of polygenic background on the penetrance of monogenic forms in Parkinson’s disease"

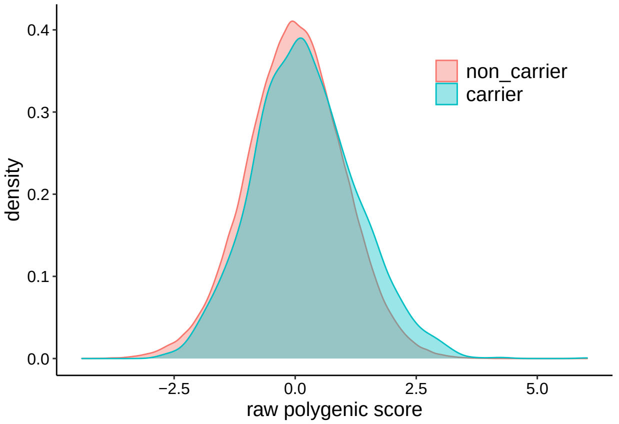


Figure S1: Distributions of the Parkinson's disease polygenic risk score (PRS) across carrier status in the UKBiobank cohort.

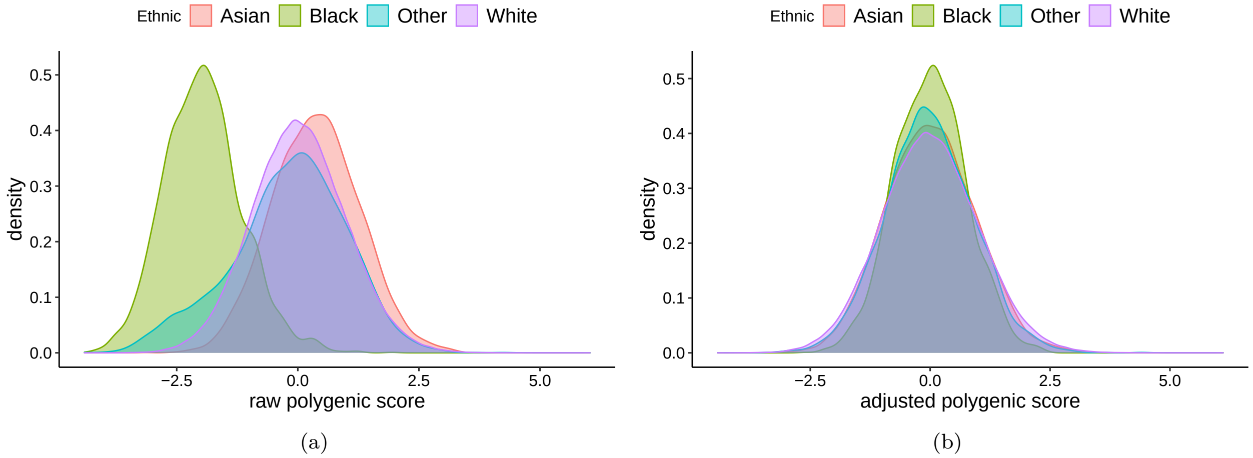


Figure S2: Distributions of the parkinson's disease (PD) PRS across different ethnic groups in the UKBiobank cohort. Raw PRS was adjusted using first four principal components. Distributions of a) raw PRS in PD. b) adjusted PRS in PD.

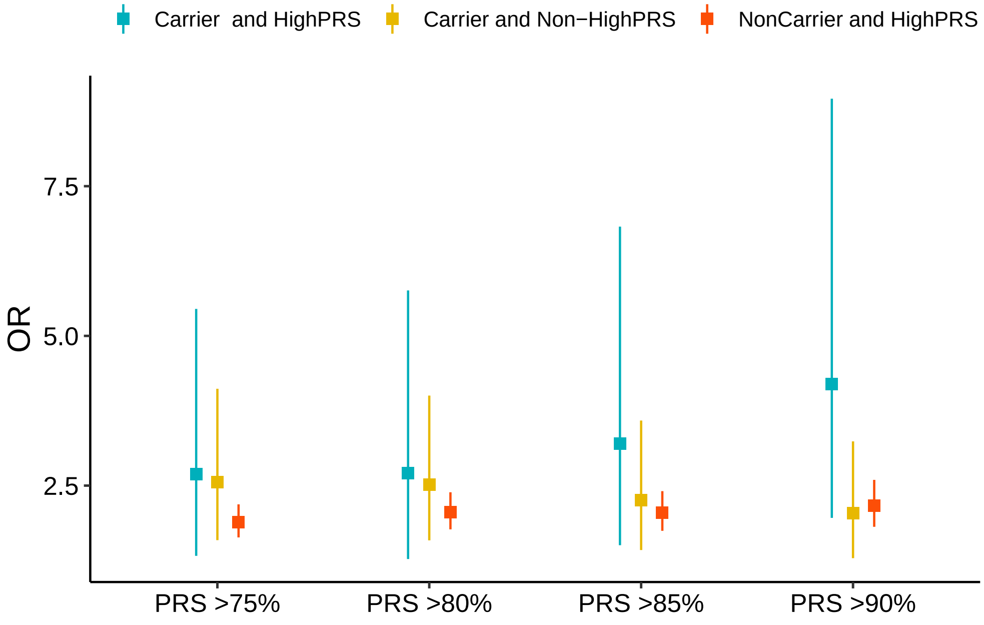


Figure S3: Impact of PRS across individuals with monogenic variants carriers.

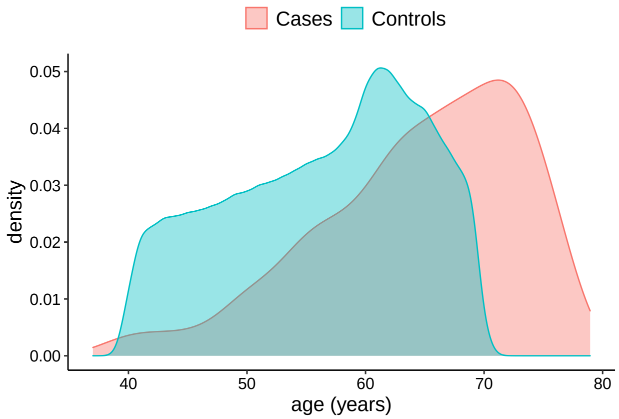


(a) Distributions of age in Cases and controls

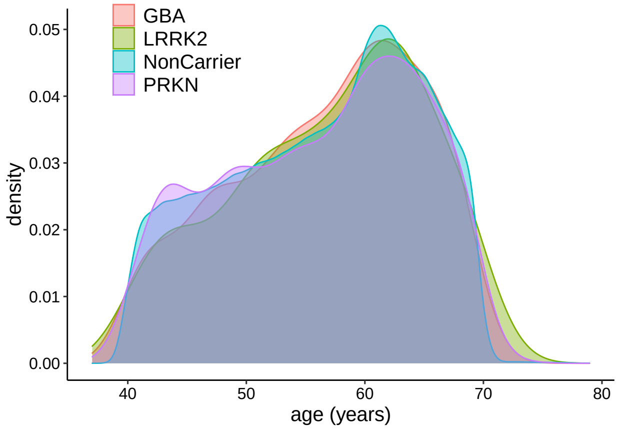


(b) Distributions of age in LRRK2, GBA, PRKN variant carriers

Figure S4: Distributions of age in the UKBiobank cohort.

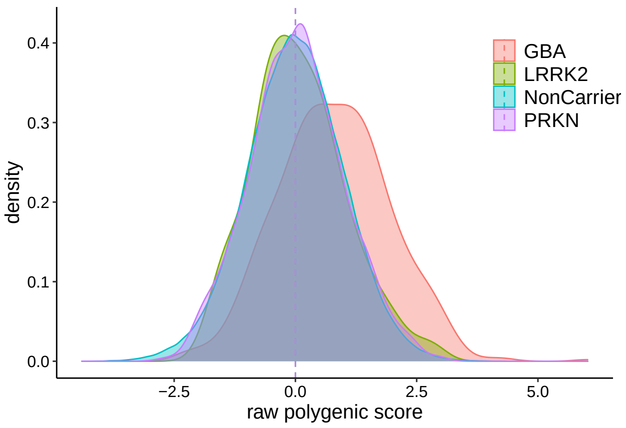


Figure S5: Distributions of the Parkinson's disease PRS across carrier of mutation in LRRK2, GBA, PRKN genes in the UKBiobank cohort.

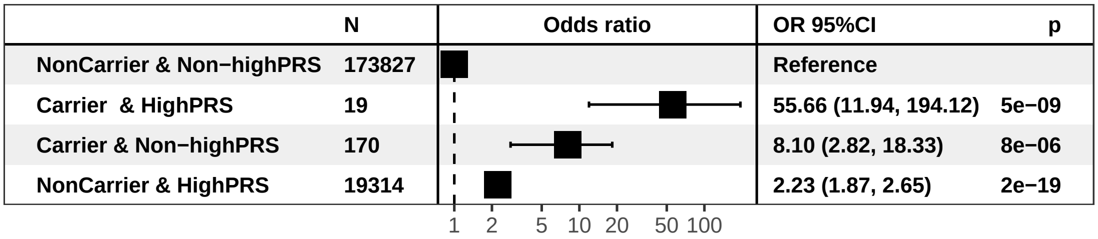


(a) LRRK2

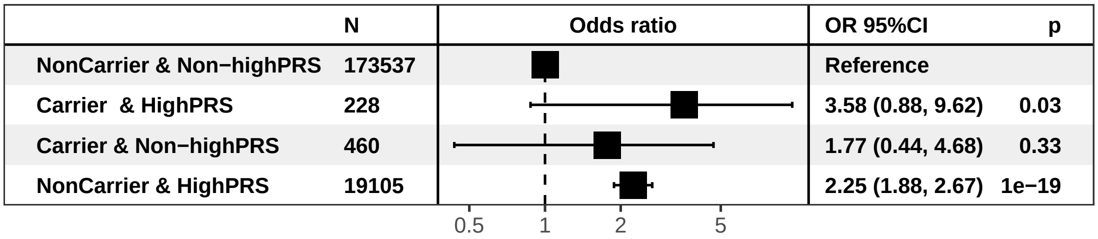


(b) GBA

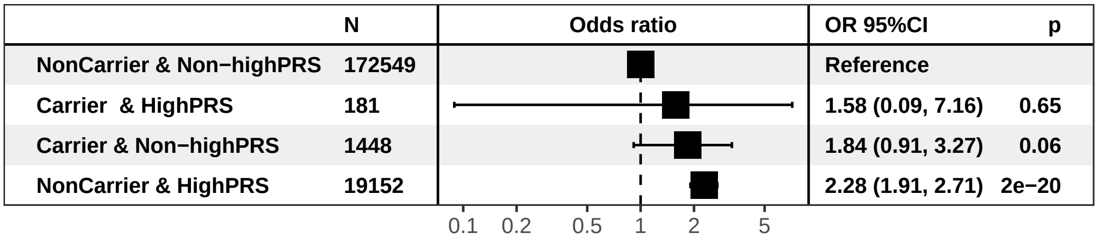


(c) PRKN

Figure S6: Impact of PRS in LRRK2, GBA, PRKN variant carriers . Carriers and noncarriers were categorised into two strata based on their PRS:Non-high (<90 % percentile), or high (>90 % percentile) risk.The odds ratio was calculated from a logistic regression model with age, sex, and the first four principal components of ancestry as covariates. The reference group was noncarriers with Nonhigh risk. The adjusted odds ratio is indicated by the black boxes. The 95% confidence intervals are indicated by the horizontal lines around the black boxes.

| Variable | | N | Odds ratio | | p |
|---|---|---|---|---|---|
| **History** | NonHistory | 185226 | | Reference | |
| | History | 8104 | | 2.61 (2.09, 3.24) | <0.001 |
| **Carrier** | NonCarrier | 190350 | | Reference | |
| | Carrier | 2980 | | 2.11 (1.38, 3.06) | <0.001 |
| **Carrier & History** | NonCarrier & NonHistory | 182434 | | Reference | |
| | Carrier & History | 188 | | 10.94 (4.83, 21.47) | <0.001 |
| | Carrier & NonHistory | 2792 | | 1.67 (1.00, 2.60) | 0.03 |
| | NonCarrier & History | 7916 | | 2.47 (1.95, 3.08) | <0.001 |

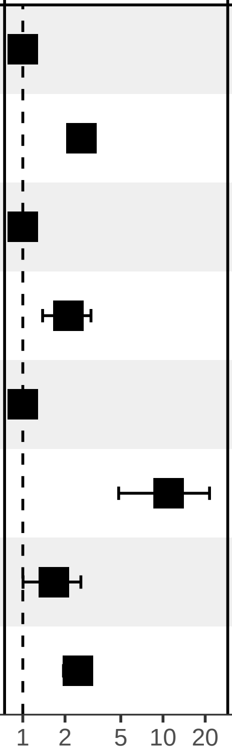


Figure S7: : Association of pathogenic carrier status and family history with risk of parkinson's disease.

Table S1: Characteristics of the 193,330 UK Biobank participants by monogenic and PRS strata.

| | Noncarrier & IntermediatePRS | Carrier & HighPRS | Carrier & LowPRS | Carrier & IntermediatePRS | Noncarrier & HighPRS | Noncarrier & LowPRS |
|---|---|---|---|---|---|---|
| Participants, N | 152397 | 474 | 239 | 2267 | 18859 | 19094 |
| Cases, N (%) | 575 (0.38) | 7 (1.48) | 2 (0.84) | 17 (0.75) | 156 (0.83) | 49 (0.26) |
| Controls, N | 151822 (99.62) | 467 (98.52) | 237 (99.16) | 2250 (99.25) | 18703 (99.17) | 19045 (99.74) |
| Male, N (%) | 68566 (44.99) | 208 (43.88) | 111 (46.44) | 1028 (45.35) | 8533 (45.25) | 8589 (44.98) |
| Female, N (%) | 83831 (55.01) | 266 (56.12) | 128 (53.56) | 1239 (54.65) | 10326 (54.75) | 10505 (55.02) |
| Age*, mean (SD) | 56.45 (8.1) | 56.06 (8.26) | 57.53 (7.75) | 56.47 (8.22) | 56.56 (8.07) | 56.51 (8.09) |
| Family history of parkinsons's disease, n (%) | 6271 (4.11) | 40 (8.44) | 9 (3.77) | 139 (6.13) | 1029 (5.46) | 616 (3.23) |

* Age: age at onset for cases, and age at recruitment for controls

Table S2: Impact of PRS across individuals with monogenic variants carriers in PPMI (Parkinson's progression markers initiative) cohort.

| | controls (%) | cases (%) |
|---|---|---|
| NonCarrier & IntermediatePRS | 81 (28.5%) | 203 (71.5%) |
| Carrier & HighPRS | 0 (0%) | 6 (100%) |
| Carrier & LowPRS | 0 (0%) | 1 (100%) |
| Carrier & IntermediatePRS | 1 (12.5%) | 7 (87.5%) |
| NonCarrier & HighPRS | 16 (17.8%) | 74 (82.2%) |
| NonCarrier & LowPRS | 47 (48.5%) | 50 (51.5%) |

Table S3: Impact of PRS across individuals with monogenic variants carriers in PPMI (Parkinson's progression markers initiative) cohort.

| history | controls (%) | cases (%) | OR (95%CI) | p-value |
|---|---|---|---|---|
| History & HighPRS | 0 (0%) | 19 (100%) | 8543959.898 (0-Inf) | 0.977 |
| History & IntermediatePRS | 4 (6.8%) | 55 (93.2%) | 7.305 (2.537-21.037) | 0.000 |
| History & lowPRS | 3 (18.8%) | 13 (81.2%) | 2.315 (0.63-8.502) | 0.206 |
| NonHistory & HighPRS | 16 (20.8%) | 61 (79.2%) | 2.029 (1.09-3.777) | 0.026 |
| NonHistory & lowPRS | 44 (53.7%) | 38 (46.3%) | 0.399 (0.236-0.676) | 0.001 |
| NonHistory & IntermediatePRS | 78 (33.5%) | 155 (66.5%) | NA | NA |
